# Serendipitous COVID-19 Vaccine-Mix in Uttar Pradesh, India: Safety and Immunogenicity Assessment of a Heterologous Regime

**DOI:** 10.1101/2021.08.06.21261716

**Authors:** Rajni Kant, Gaurav Dwivedi, Kamran Zaman, Rima R Sahay, Gajanan Sapkal, Himanshu Kaushal, Dimpal A. Nyayanit, Pragya D Yadav, Gururaj Deshpande, Rajeev Singh, Sandeep Chaowdhary, Nivedita Gupta, Sanjay Kumar, Priya Abraham, Samiran Panda, Balram Bhargava

**Affiliations:** Indian Council of Medical Research-Regional Medical Research Centre (RMRC), Gorakhpur, Uttar Pradesh, India, Pin-273013; Indian Council of Medical Research-National Institute of Virology (ICMR-NIV), Pune, Maharashtra, India, Pin-411021; Chief Medical Officer, Community Health Centre, Siddarthnagar, Uttar Pradesh, India, Pin-272207; Indian Council of Medical Research, V. Ramalingaswami Bhawan, P.O. Box No. 4911, Ansari Nagar, New Delhi, India Pin-110029; Command Hospital (Southern Command), Armed Forces Medical College, Pune, Maharashtra, India Pin-411040

## Abstract

Immunization program against COVID-19 in India started with two vaccines; AstraZeneca’s ChAdOx1-nCov-19 (termed Covishield in India) and inactivated whole virion BBV152 (Covaxin); homologous prime-boost approach was followed. However, eighteen individuals, under the national program, inadvertently received Covishield as the first jab and Covaxin as the second. We compared the safety and immunogenicity profile of them against that of individuals receiving either Covishield or Covaxin (n=40 in each group). Lower and similar adverse events following immunization in all three groups underlined the safety of the combination vaccine-regime. Immunogenicity profile against Alpha, Beta and Delta variants in heterologous group was superior; IgG antibody and neutralising antibody response of the participants was also significantly higher compared to that in the homologous groups. The findings suggest that immunization with a combination of an adenovirus vector platform-based vaccine followed by an inactivated whole virus vaccine was not only safe but also elicited better immunogenicity.

## Introduction

Vaccination against COVID-19 in India was rolled out on January 16th, 2021; about a year after the detection of Severe Acute Respiratory Syndrome Coronavirus 2 (SARS-CoV-2) in the country^1^. Two vaccines, namely Covaxin and Covishield were the cornerstones of this program, which was hailed as the world’s largest vaccination campaign against the COVID-19 pandemic^2,3,4,5^. While Covishield is the in-country version of AstraZeneca vaccine manufactured by Serum Institute of India, Covaxin is an inactivated whole virus vaccine (BBV152) developed jointly by Indian Council of Medical Research-National Institute of Virology (ICMR-NIV) and Bharat Biotech International Limited (BBIL). The regime for both the vaccines initially included a second homologous booster dose following a priming dose at an interval of 4 weeks. With gradually emerging evidence, the gap between the two doses of Covishield was increased to 6-8 weeks and later to 12 weeks^6,7,8^. India prioritized frontline workers including healthcare professionals, elderly above the age of 60 and adults with co-morbidity for vaccination before reaching out to all adults. Such targeted approach with its potential to flatten the curve of symptomatic infections in the country and to reduce the mortality was considered as a smart public health approach^9^.

During expansion of the aforementioned vaccination program, a group of individuals in the northern State of Uttar Pradesh in India received Covishield as the first dose followed by inadvertent administration of Covaxin as the second dose at an interval of six weeks (n=20). The nationwide vaccination program at this time entered into 4 months of its existence and the event of mixed dosing raised considerable anxiety in public domain with a potential to contributing to vaccine hesitancy. We conducted the current investigation against this backdrop.

The article presents data on reactogenicity and immunogenicity in individuals who received the heterologous vaccine regime described above. To the best of our knowledge, this is the first study, which reports the effects of heterologous prime-boost vaccination with an Adenovirus vectored vaccine followed by an inactivated whole virus vaccine. The results of this investigation were compared against the ones generated by two homologous prime-boost vaccination scenario; Covishield-Covishield (CS) or Covaxin-Covaxin (CV) combination as the first and second dose respectively, which is the ongoing program reality in India.

## Result

A total of 18 participants were in the heterologous group (two participants were unwilling and were excluded). Among them, 11 were male (M/F: 11/7) with a median age of 62 years (IQR 54.25-69.75). A comorbid condition (hypertension) was reported in one (5.5%) individual. In both the homologous groups, 40 individuals were included. In the CS group, 22 were males (M/F: 22/18) with a median age of 65.5 years (IQR 62-69) and co morbidity was present in 4 (10%) individuals. In the CV group, 23 were females (M/F:17/23) with a median age of 56 years (IQR 45.5-63) and 3 (7.5%) had co-morbidities (Table-1) (Figure-1).

**Table-1:** Clinico-demographic profile of participants: Demographic and clinical characteristics of the study population. Demographic characteristics and vaccine-related data are shown for the three vaccination regimens [homologous Covishield vaccination (CS, n=40), homologous Covaxin vaccination (CV, n = 40) and heterologous Covishield plus Covaxin vaccination (CS/CV, n=18). In addition, information on lymphocyte subpopulations is also provided

| 1<sup>st</sup> vaccine +<br>2<sup>nd</sup> vaccine | Covishield +<br>Covishield<br>(CS) | Covaxin + Covaxin (CV) | Covishield +<br>Covaxin<br>(CS/CV) | p-value |
| --- | --- | --- | --- | --- |
| **Total vaccines enrolled** | n=40 | n=40 | n=18 |  |
| **Median age (IQR)** | 65.5 [62-69] | 56 [45.5-63] | 62[54.25-69.75] | <0.001<sup>\$</sup> |
| **Male** | 22 (55%) | 17 (42.5%) | 11 (61.11%) | 0.341* |
| **Female** | 18 (45%) | 23 (57.5%) | 7 (38.89%) |  |
| **Co-morbidities** | 4 (10%) | 3 (7.5%) | 1 (5.55%) | 0.83* |
| **Type of comorbidities** | Hypertension, Diabetes, Chronic Heart Disease and Obesity | Hypertension/Diabetes, Obesity, Chronic Liver disease | Hypertension |  |
| **% Lymphocytes median (IQR)** | 23.19 (19.73 - 26.98) | 27.71 (24.03 - 32.09) | 28.63 (21.35 - 32.50) | 0.0519<sup>£</sup> |
| **% CD4/CD3+ T-cells, median (IQR)** | 43.42 (35.35 - 54.30) | 51.26 (41.03 - 59.41) | 43.85 (35.61 - 58.28) | 0.0796<sup>£</sup> |
| **% CD3 T-cells/lympho median (IQR)** | 57.75 (48.61 - 63.75) | 56.36 (44.33 - 63.58) | 51.46 (44.18 - 60.97) | 0.525<sup>£</sup> |
| **% CD8/CD3+ T-cells, median (IQR)** | 47.66 (40.03 - 55.02) | 39.4 (33.87 - 49.27) | 46.45 (36.66 - 54.52) | 0.0329<sup>£</sup> |
| **% CD19 B-cells/ CD45+ cells, median (IQR)** | 11.86 (8.678 - 15.15) | 10.61 (8.393 - 13.94) | 11.2 (8.245 - 14.80) | 0.5223<sup>£</sup> |
| **% CD16+56 NK cells/CD45+ cells, median (IQR)** | 18.55 (14.40 - 27.65) | 19.44 (12.06 - 27.35) | 19.11 (12.25 - 27.53) | 0.9497<sup>£</sup> |

**Figure 1:**
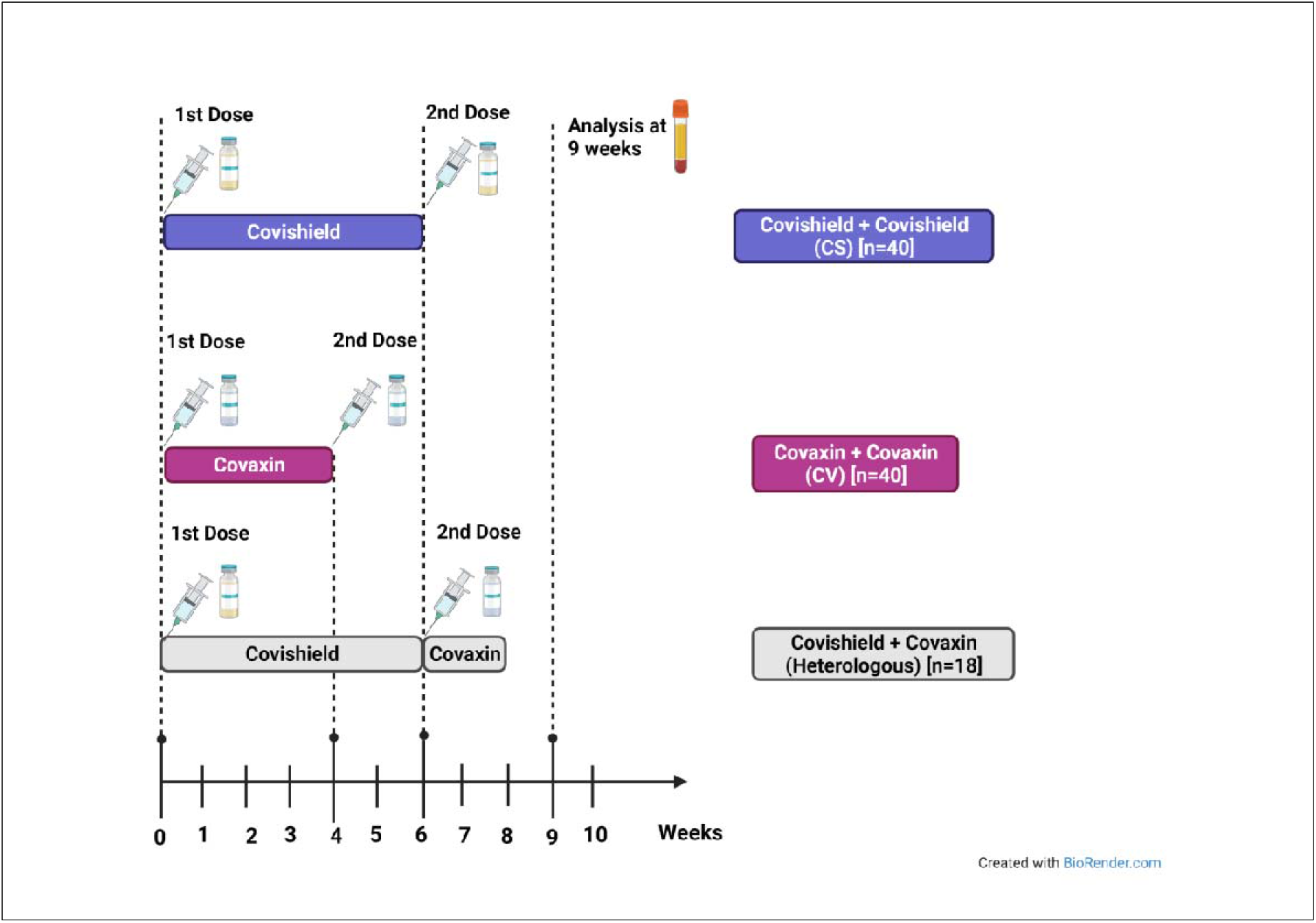
Study design

Reactogenicity analysis was carried out based on solicited local and systemic AEFIs reported in the three groups within seven days of immunisation. None of the participants enrolled in the study had any serious AEFI within 30 minutes of immunisation with the first or second dose. The most common local AEFI reported after first and second dose was pain at injection site; CS group [n=40 (1^st^ dose 5%; 2^nd^ dose 5%)], CV group [n=40, (1^st^ dose 7.5%; 2^nd^ dose 7.5%)] and heterologous group [n=18, (1^st^ dose 11.1%; 2^nd^ dose nil)]. No other local AEFI such as erythema, induration, pruritis or pustule formation was recorded by any of the participants. Most commonly reported systemic AEFI were pyrexia and malaise. The frequency of pyrexia reported was: CS group [n=40, (1^st^ dose 20%; (2^nd^ dose 15%)], CV group [n=40, (1^st^ dose 30%; 2^nd^ dose 15%)] and heterologous group [n=18, (1^st^ dose 27.77%; 2^nd^ dose 11.1%)]. Malaise was reported in CS group [n=40, (1^st^ dose 5%; 2^nd^ dose 5%)], CV group [n=40, (1^st^ dose 32.5%; 2^nd^ dose 15%) and heterologous group [n=18, (1^st^ dose 33.3%, 2^nd^ dose 5.5%)]. No other systemic AEFI like, urticaria, nausea, vomiting, arthralgia or cough was reported. The pyrexia was of low to moderate grade and was managed by administration of paracetamol and subsided in all participants within 3 to 4 days post vaccination. The frequency of consumption of paracetamol was; CS group [n=40, (1^st^ dose 12.5%; 2^nd^ dose 7.5%)], CV group [n=40, (1^st^ dose 11.1%; 2^nd^ dose 17.5%)] and heterologous group [n=18, (1^st^ dose 11.1%; 2^nd^ dose nil)] (Figure 2a-d).

**Figure-2:**
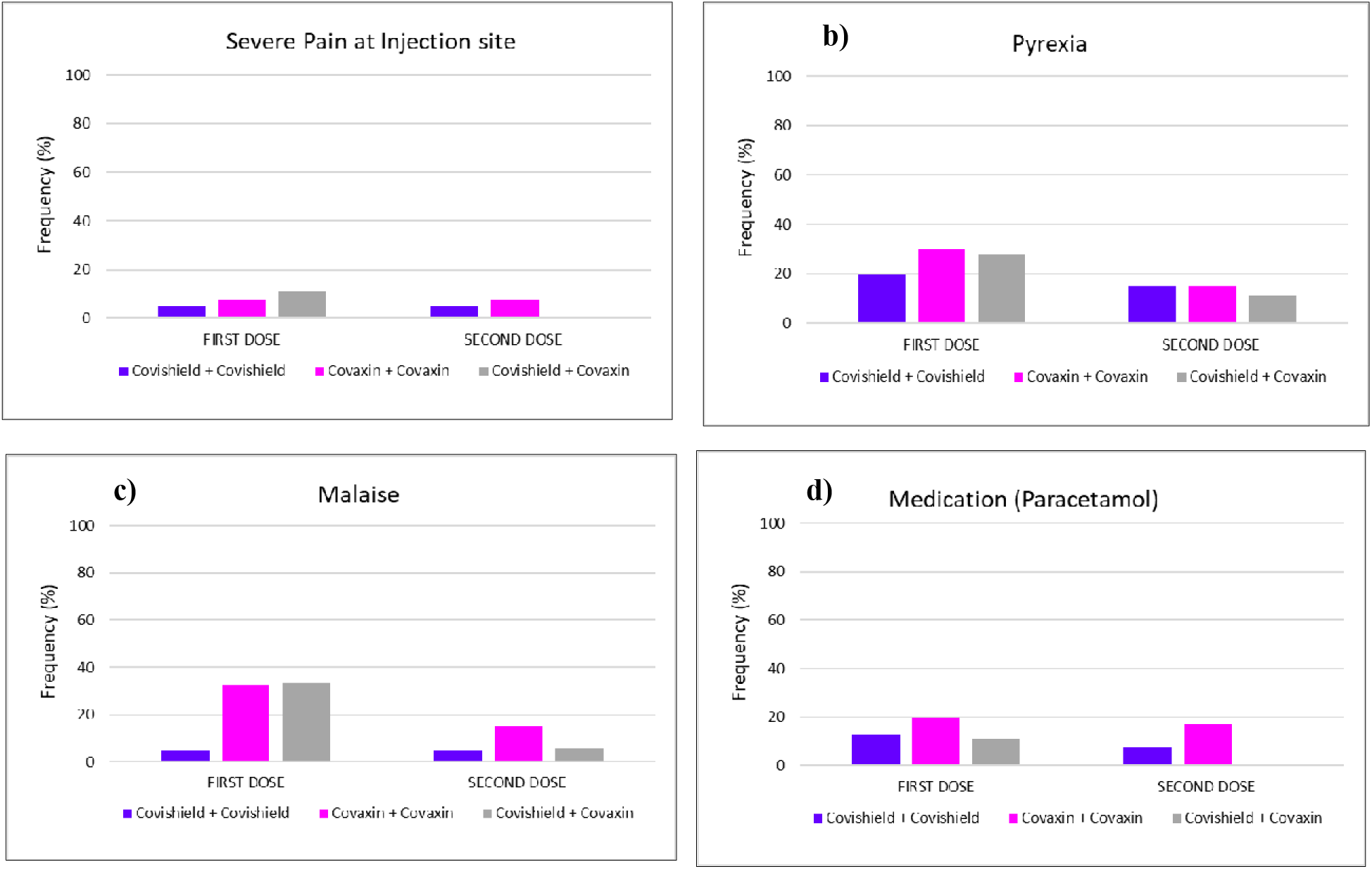
Frequency of solicited local and systemic adverse events in homologous Covishield, homologous Covaxin and heterologous groups. a) Severe pain at injection site b) Pyrexia, c) Malaise and d) consumption of medication (Paracetamol).

The frequency of local and systemic AEFI was lower after the second dose in all the participants of the three groups. CV group participants had higher frequency of AEFI than CS group. Overall, the frequency of AEFI in the heterologous group was similar to the participants of the CV/CS groups establishing the safety of heterologous vaccination.

The ELISA titres against S1-RBD, N protein and inactivated SARS-CoV-2 (whole virus) was compared in sera of the three groups and analysed statistically using the Kruskal-Wallis test along with Dunns’ multiple comparisons. The geometric mean (GM) of the ELISA for participants from the CS, CV and heterologous groups against S1-RBD was 2260 (95% CI: 1881-2716), 710 (95% CI: 461-1092) and 1866 (95% CI: 1003-3472) respectively. The GM of N-protein in CS, CV and heterologous groups were 353.7(95% CI: 219.9-568.9), 742.4(95% CI: 485.8-1134) and 1145(95% CI: 520.7-2520) respectively. Finally, GM of IgG titres against inactivated SARS-CoV-2 (whole virus) in sera of the participants of CV, CS and heterologous groups were 111 (95% CI: 98.59-124.9), 86(95% CI: 138.2-252.0), 171.4(95% CI: 121.3-242.3) respectively (Figure 3a-c). These findings reveal a significantly higher IgG antibody response of participants of the heterologous group as compared to either CS or CV group.

**Figure-3:**
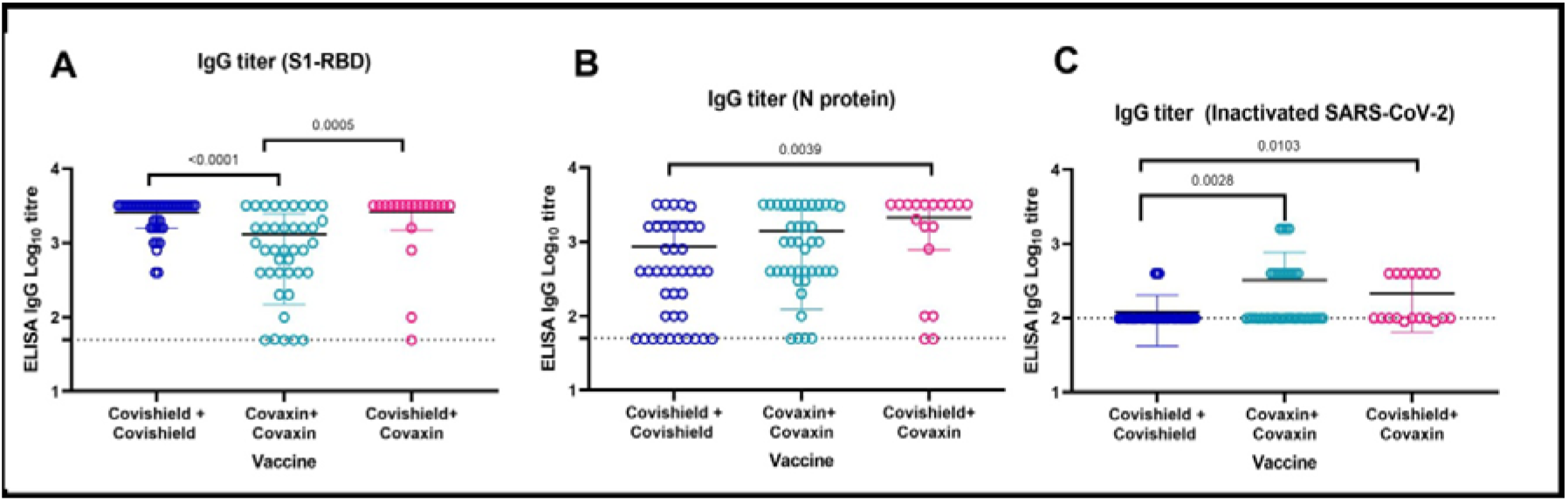
ELISA titre of individual sera vaccinated with the different vaccines. **a)** Anti-SARS-CoV-2 IgG titres of vaccinated individual’s sera for S1-RBD protein for homologous Covishield [n=40, (blue)], homologous Covaxin [n=40, (green)] and heterologous Covishield/Covaxin[n=18, (pink)]. **b)** N-specific IgG level in serum in the three groups. **c)** Inactivated SARS-CoV-2 IgG levels in serum of the three groups. The statistical significance was assessed using a two-tailed Kruskal Wallis test with Dunn’s test of multiple comparisons; p-value less than 0.05 were considered to be statistically significant. The dotted line on the figures indicates the limit of detection of the assay. Data are presented as mean values +/− standard deviation (SD).

The PRNT50 assay was performed with the sera of participants of all the three groups against B.1, Alpha, Beta and Delta strains. The geometric mean titre (GMT) with 95% confidence interval (CI) for the CS group against the B.1, Alpha, Beta and Delta strain was 162(95% CI: 76.74-342), 122.7(95% CI: 59.36-253.7), 48.43(95% CI: 19.71-119) and 51.99(95% CI: 19.65-137.6) respectively. The GMT for the CV group against the B.1, Alpha, Beta and Delta strain was 156.6(95% CI: 105.2-233.1), 112.4(95% CI: 76.56-164.9), 52.09(95% CI: 34.9-77.73) and 54.37(95% CI: 27.26-108.4) respectively. The GMT for the heterologous group against B.1, Alpha, Beta and Delta strains was 539.4(95% CI: 263.9-1103), 396.1(95% CI: 199.1-788), 151(95% CI: 80.21-284.3) and 241.2(95% CI: 74.99-775.9) respectively. Wilcoxon signed-ranks test was performed to analyse the statistical significance of the neutralization titre observed for the participants of the heterologous group.

The GMT ratio was higher for the Beta strain relative to B.1 strain [3.35(95% CI: 2.87-3.89; p-value =0.0062)] in comparison to Alpha [1.32(95% CI: 1.29-1.35); p-value >0.999] and Delta [3.12(95% CI: 2.49-3.91); p-value=0.0820] in the participants of CS group (Figure 4a). Similarly, the GMT ratio was higher for the Beta strain relative to B.1 strain [3.01 (95% CI: 3.00-3.01; p-value =0.0001)] in comparison to Alpha [1.39 (95% CI: 1.37-1.41); p-value >0.999] and Delta [2.88 (95% CI: 2.15-3.86); p-value=0.0196] in the participants of CV group (Figure 4 b). The GMT ratio was higher for the Beta strain relative to B.1 strain [3.57(95% CI: 3.29-3.88; p-value =0.0125)] in comparison to Alpha [1.36(95% CI: 1.33-1.40); p-value >0.999] and Delta [GMT ratio: 2.24(95% CI: 1.42-3.52); p-value >0.999] in the patients receiving heterologous vaccine (Figure 4 c). The analysis of the PRNT50 data with all the four strains shows that higher NAbs were observed against the Alpha strain in the sera of immunised participants of all the three groups followed by the Delta strain.

**Figure-4:**
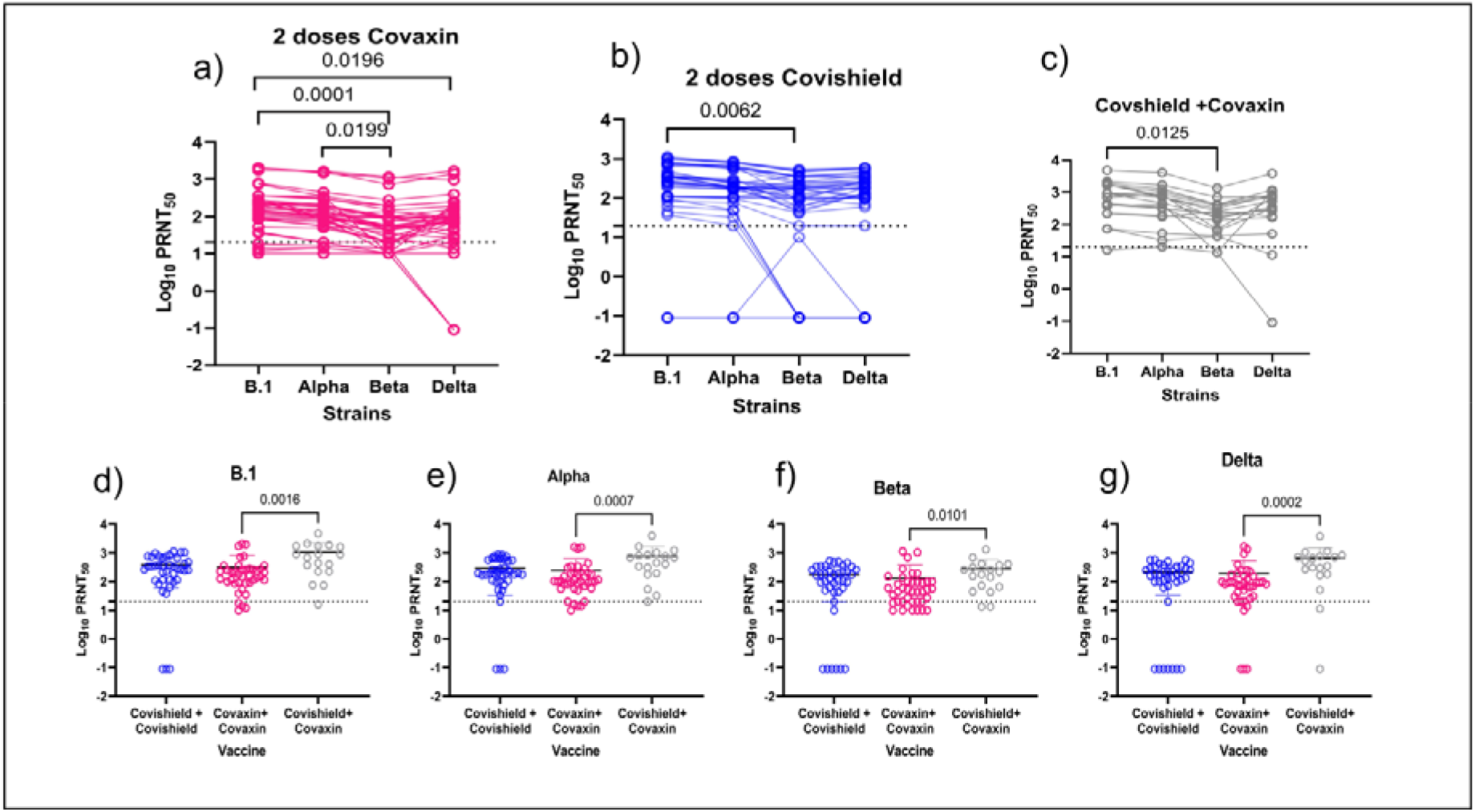
Neutralization of individual sera treated with different vaccines against B.1, alpha, beta and delta strains: NAb titre of individual sera against the B.1, beta, alpha and delta strain administered with 2 doses of Covishield (n=40) (a), 2 doses of Covaxin (n=40)(b) and a single dose of Covishield followed by Covaxin (n=18) (c). A matched pair two-tailed pair-wise comparison was performed using the Wilcoxon signed-rank test to analyze the statistical significance. Neutralization titre of individual’s sera vaccinated against different strains; Alpha (d), Beta (e), Delta (f), and B.1 (g). The statistical significance was assessed using a two-tailed Kruskal Wallis test with Dunn’s test of multiple comparisons was performed to analyze the statistical significance. A p-value less than 0.05 were considered to be statistically significant for the comparison. The dotted line on the figures indicates the limit of detection of the assay. Data are presented as mean values +/− standard deviation (SD).

A significant difference in NAb titres of the sera obtained from the participants of CV group relative to sera of heterologous group against each strain was observed (Figure 4 d-g). Further, it was also observed that participants of the heterologous group had ~3-fold higher titre in comparison to CS and CV groups (Figure 4 d-g). Kruskal-Wallis test along with Dunns’ multiple comparisons was performed to assess the significance in NAb titres of the sera obtained from immunised participants against different strains.

The anti-SARS-CoV-2 IgG titers of vaccinated individual demonstrated the higher neutralizing titer for the S1-RBD protein followed by N protein (Figure 5a-c). Neutralization of individual sera, with average age ranging approximately between 62.0-63.0 yrs, administered with different vaccines against B.1, Alpha, Beta and Delta variants were compared to observe the effect of age on Nab titers. It is observed that the neutralizing NAbs were 1.25, 3.95 and 1.30 fold reduced in heterologous group for Alpha, Beta and Delta respective compared to B.1 strain. Similarly the NAb were reduced homologous Covishield [1.33, 3.9, and 2.74], homologous Covaxin [1.4, 2.45, and 2.08] for Alpha, Beta and Delta respective compared to B.1 strain (Figure-5 d-f).

**Figure 5:**
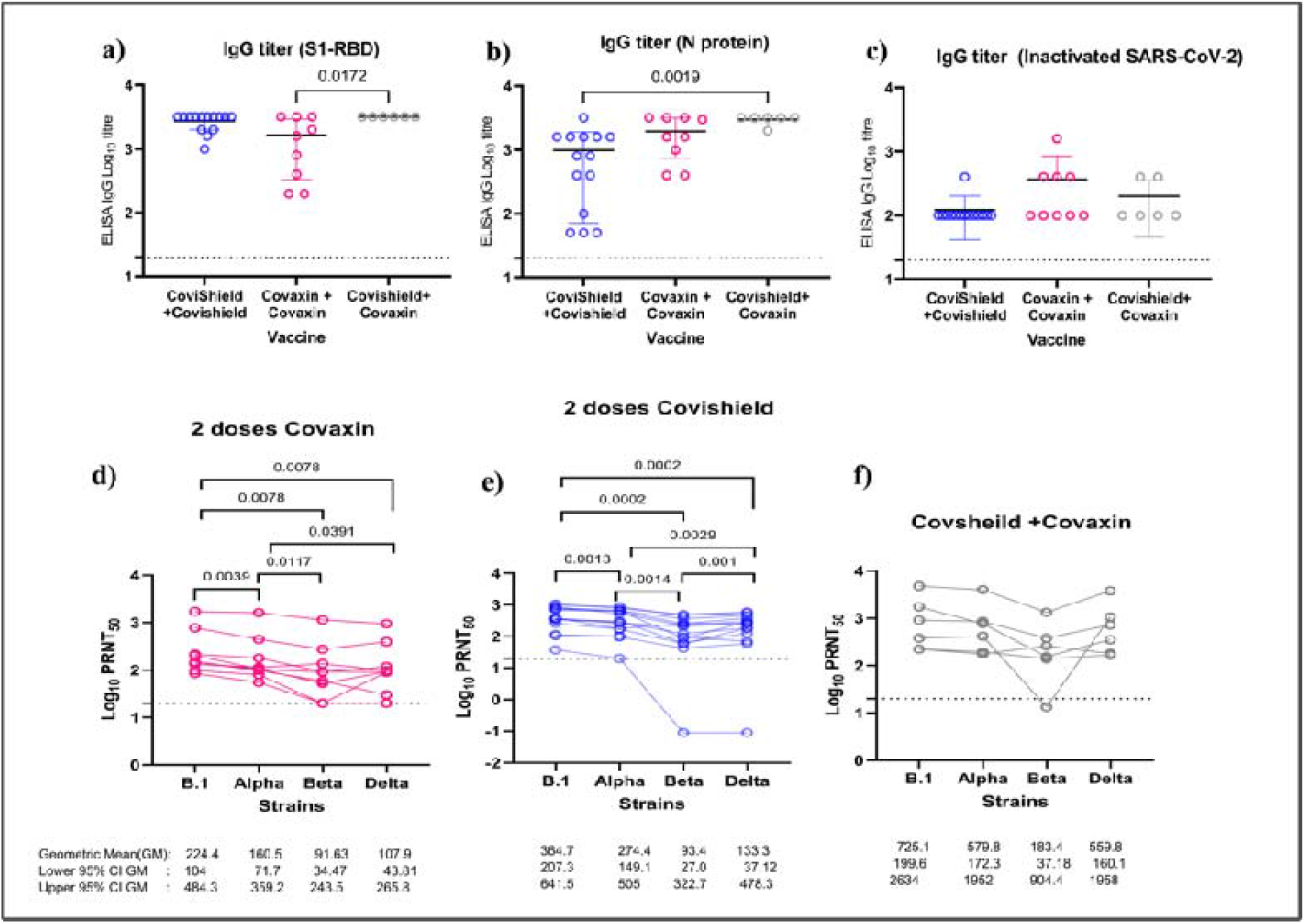
ELISA and the neutralization titer of individual sera, with average age ranging approximately between 62.0-63.0 yrs, administered with different vaccines against B.1, Alpha, Beta and Delta strains: **a)** Anti-SARS-CoV-2 IgG titres of vaccinated individual’s sera for S1-RBD protein **b)** N-specific IgG level and **c)** Inactivated SARS-CoV-2 IgG levels in serum for 2 doses of Covishield (n=14), 2 doses of Covaxin (n=9) and a single dose of Covishield followed by Covaxin (n=6). The statistical significance was assessed using a two-tailed Kruskal Wallis test with Dunn’s test of multiple comparisons. NAb titre of individual sera against the B.1, Beta, Alpha and Delta strain administered with 2 doses of Covishield (d), 2 doses of Covaxin(d) and a single dose of Covishield followed by Covaxin (f). A matched pair two-tailed pair-wise comparison was performed for the comparing the individual sera administered with 2 doses of Covaxin while the rest were compared using the Wilcoxon signed-rank test to analyse the statistical significance. A p-value less than 0.05 were considered to be statistically significant for the comparison. The dotted line on the figures indicates the limit of detection of the assay. Data are presented as mean values +/− standard deviation (SD).

Assessment based on the surface marker of the gated lymphocytes from lysed whole blood (Figure 6) was found to be comparable among all the study groups i.e., heterologous vaccinated group and CS or CV vaccinated groups in terms of the percentage of cells expressing CD3, CD4, CD16+56 or CD19. However, % CD8+ T cells in the CS group (median (IQR): 47.66 (40.03–55.02), p < 0.05) was found significantly high compared to CV group (median (IQR): 39.40 (33.87– 49.27) (Figure 7a-f).

**Figure-6.**
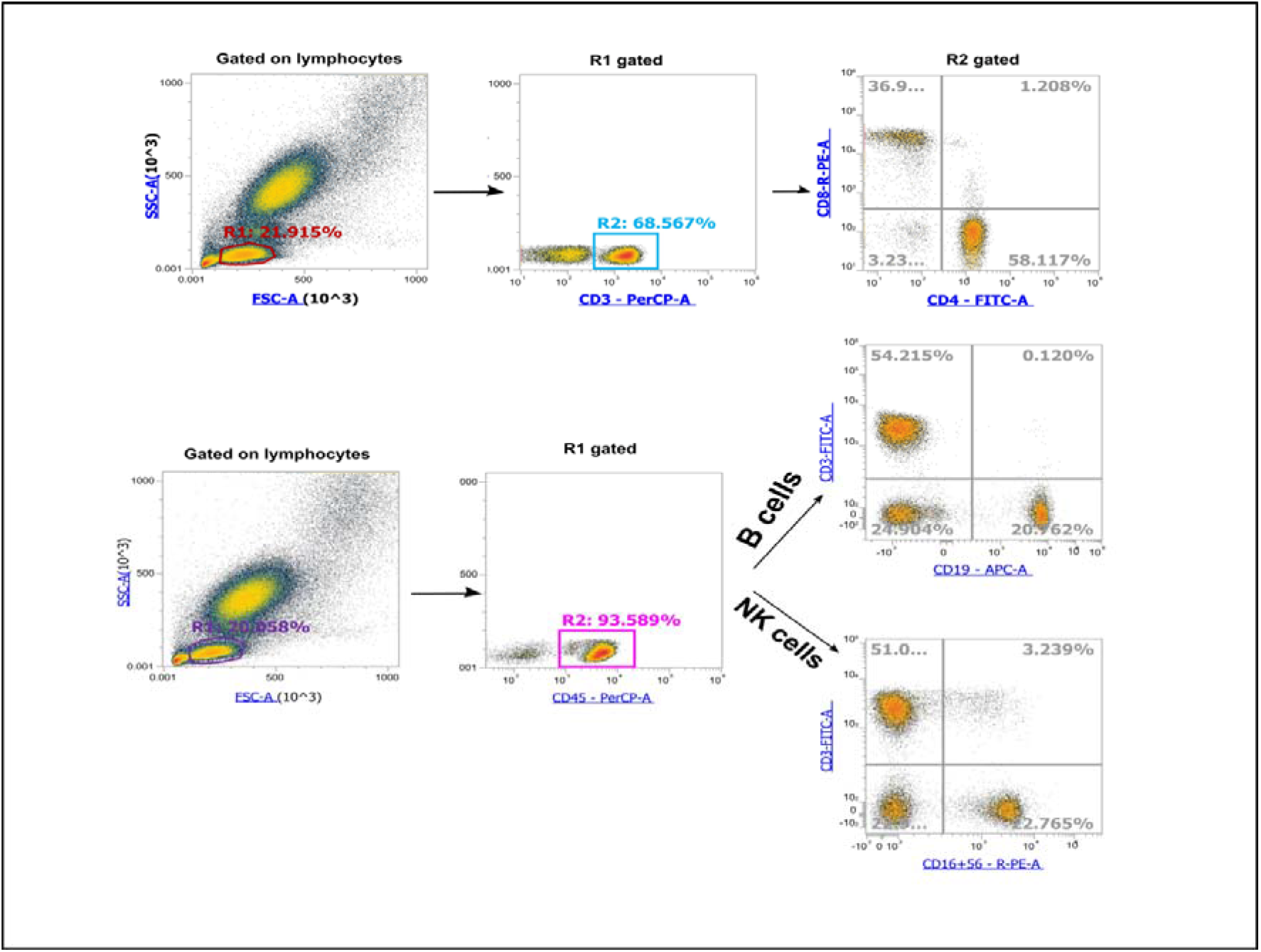
Gating strategy for phenotyping of lymphocyte subsets in whole blood. First, lymphocytes were gated based on forward- and side-scatter; CD3+ and CD45+ cells were gated on lymphocytes. CD4+ and CD8+ cells were gated on CD3+ cells whereas CD19+ B cells and CD16+56+ NK cells were gated on CD45+ cells.

**Figure-7.**
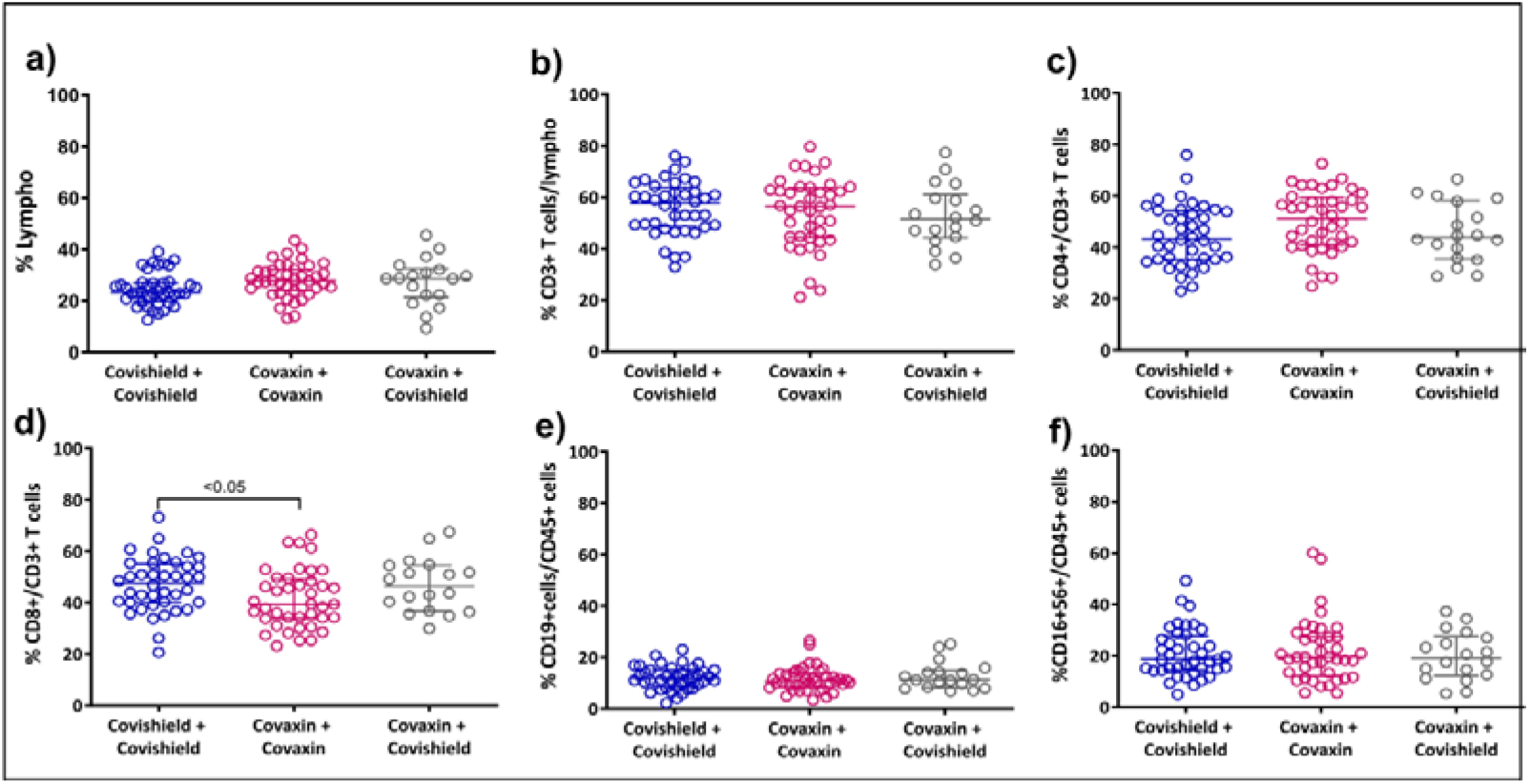
Flow cytometry analysis of lymphocyte subsets in COVID-19 vaccinated groups. Whole blood samples from a single dose of Covishield followed by Covaxin (n=18), 2 doses of Covishield (n=40) and 2 doses of Covaxin (n=40) were surface-stained with appropriate antibodies and analyzed on Attune NxT flow cytometry using Attune NxT software. The figures represent % lymphocytes (a) % CD3+ T cells/lymphocytes (b), % of CD4+ (c) and CD8+ cells (d) of total CD3+ T cells. Similarly, % CD19+ B cells (e) and % CD16+56+ cells (f) were gated on CD45+ cells. The data were analyzed using GraphPad Prism 9.0 (GraphPad Software, San Diego, CA, USA). Statistical significance was determined by the non-parametric Kruskal–Wallis test, followed by the post-hoc Dunn’s multiple comparison test for more than two groups. The groups that were found significant using the Kruskal–Wallis test, were further analyzed using the Mann–Whitney test. The statistical tests were two-tailed and P values <0.05 were considered significant. Horizontal lines indicate median values with an interquartile range.

## Discussion

Earlier studies have reported the reactogenicity and immunogenicity of mixed vaccination regimens. Shaw *et.al* reported increase in proportion of participants with fever, higher paracetamol usage and increased systemic reactogenicity in a group of individuals primed with ChAdOx1 and followed by BNT162b2 for boost immunisation^10^. Hillus *et.al*, showed no major differences in local or systemic reactogenicity amongst the recruited health care workers who were given heterologous ChAdOx1/BNT162b2 vaccination in comparison to homologous BNT162b2/BNT162b2 vaccination^11^. In our study, no major systemic AEFIs were reported and reactogenicity profile of the participants of heterologous group was comparable to homologous CS and CV groups. The profile of solicited AEFIs in the heterologous group was similar to that of either Covaxin^2,3^ or Covishield^12^ Despite the high median age of the participants of the heterologous group (62 years) in our study, the reactogenicity profile demonstrated that mixing of the two vaccines based on different platforms is safe.

Spencer *et al*. have shown that the heterologous vaccination regimens elicit better antibody responses post-vaccination with good NAb titres after heterologous prime-boost compared to the homologous vaccination with ChAdOx1 nCoV-19, which is consistent with previous data in mice, pigs and non-human primates shown by Graham *et al* and Doremalen *et al* ^13,14,15^. Our study also demonstrated that the NAb titers in the heterologous group were significantly higher as compared to the homologous groups of CS and CV. The sera of the heterologous group was efficient in neutralizing VOCs; Alpha, Beta and Delta as we have previously reported with two doses of Covaxin and Covishield^16. 17,18^.

The humoral immune response in the heterologous group in our study (IgG titers for S1-RBD, N protein and inactivated SARS-CoV-2 antigen) was significantly higher in comparison to the homologous groups (CS and CV). Similar findings have been reported by other studies of heterologous vaccines carried out recently^19–22^.

The phenotype and proportion of different peripheral blood lymphoid subsets were investigated to understand generalized CMI in different cohorts of individuals immunised in the three groups. The study demonstrated a comparable proportion of the major lymphoid cells such as B-lymphocytes, Natural Killer cells, CD3+ T cells in the peripheral blood among heterologous and CS or CV groups. This is suggestive of no major changes in the generalized cellular immunity among the vaccinated study groups. However, there was a significant increase in the percentage of CD8+ T cells in the CS cohort group compared to the CV group, indicative of elevated cytotoxic T activity in the Covishield group (Figure-7a-f).

There are a few limitations in our study. The sample size of 18 participants is small, the follow up period is only 60-70 days after immunisation with the first dose and baseline serological and immunological data of the participants is not available. However, there are several major strengths in this study. This is the first report of heterologous immunisation with an adenovirus vector based and an inactivated whole virion vaccine in humans demonstrating safety and significantly improved immunogenicity. Qian He *et al*., had earlier reported similar findings in a mouse model^9^. Comparable proportion of solicited AEFIs in the heterologous and homologous groups despite the elderly age group (mean age 62 years) demonstrates the safety of combination regimen. Immunogenicity profile studied against the VOCs; Alpha, Beta and Delta variants demonstrates significantly higher titers in the heterologous group.

Overall, this study demonstrates that immunization with a heterologous combination of an adenovirus vector platform-based vaccine followed by an inactivated whole virus vaccine is safe and elicits better immunogenicity than two doses of homologous vaccination, using the same vaccines. These findings have an important implication for the COVID-19 vaccination program wherein heterologous immunisation will pave the way for induction of improved and better protection against the variant strains of SARS-CoV-2. Such mixed regimens will also help to overcome the challenges of shortfall of particular vaccines and remove hesitancy around vaccines in people’s mind that could have genesis in programmatic ‘errors’ especially in settings where multiple COVID-19 vaccines are being used. However, to conclusively prove these findings a multicentre RCT needs to be carried out.

## Methods

Inadvertent vaccine interchangeability with 1^st^ dose of Covishield and 2^nd^ of Covaxin was reported under program setting in twenty individuals in Audai Kalan village of Siddharthnagar district of eastern Uttar Pradesh, India. This gave us an opportunity to study the safety and immunogenicity of the heterologous combination to understand the potential benefits of such an approach.

### Ethical statement

The study was approved by the Institutional Ethics Committee of the ICMR-Regional Medical Research Centre (ICMR-RMRC), Gorakhpur (IHEC Number-RMRCGKP/EC/2021/2.1). Written and informed consent was obtained from all the participants enrolled in the study before the collection of clinical data and samples.

### Enrolment of the participants

A total of 98 vaccine recipients having completed a period of two weeks or more after the second dose were included. The study duration was from May to June 2021. Participants were recruited in three cohorts: heterologous group (n=18, first dose Covishield, second dose Covaxin administered at an interval of six weeks), Covishield group [(CS) (n = 40, two doses given six weeks apart] and Covaxin group [(CV) (n = 40, two doses administered four weeks apart]. (Figure 1).

### Data and sample collection

Information on demographic profile, vaccination history, clinical history including recent illness, co-morbidities and details of adverse events following immunization (AEFIs) were recorded by trained investigators of the ongoing national vaccination programme against SARS-CoV-2 of India. The filled in paper-based forms were utilised for selection of participants and data collection. Further, data was also collected by home visits and personal interviews as well as telephonic interactions. All data was entered in the predesigned questionnaire developed for the study. Inclusion criteria was defined as individuals above 18 years of age of either sex eligible to receive COVID-19 vaccines in the national immunisation program of India. Study participants selected for reactogenicity and immunogenicity analysis were matched for sex and age in the three groups. 5 ml venous blood was collected in serum separator gel tube vacutainers from each of the participants 60-70 days after receipt of the first dose. Venous blood was centrifuged (3000 rpm for 10 min), serum was separated and was shipped to ICMR-NIV, Pune as per the existing International Air Transport Association guidelines^23^. The samples were processed for anti-SARS-CoV-2 IgG against S1 region of the RBD (S1-RBD), Nucleocapsid (N) protein, inactivated whole SARS-CoV-2 antigen, neutralizing antibody (NAb) response and phenotyping of lymphoid cells of lysed whole blood.

### Anti-SARS-CoV-2 IgG antibody evaluation

We evaluated anti-SARS-CoV-2 IgG antibody response against S1-RBD, N-protein and the whole antigen. Briefly, 96-well ELISA plates (Nunc, Germany) were coated with SARS-CoV-2 specific antigens (S1-RBD at a concentration of 1.5µg/well and N protein at a concentration of 0.5µg/well in PBS pH 7.4). The plates were blocked with a Liquid Plate Sealer (CANDOR Bioscience GmbH, Germany) and Stabilcoat (Surmodics) for two hours at 37°C. The plates were washed twice with 10 mM phosphate buffer saline (PBS), pH 7.4 with 0.1 per cent Tween-20 (PBST) (Sigma-Aldrich, USA). The sera were serially diluted four-fold and added to antigen-coated plates and incubated at 37°C for one hour. These wells were washed five times using 1× PBST and followed by addition 50 μl/well of anti-human IgG horse radish peroxidase (HRP) (Sigma) diluted in Stabilzyme Noble (Surmodics). The plates were incubated for half an hour at 37°C and then washed as described above. Further, 100 μl of TMB substrate was added and incubated for 10 min. The reaction was stopped by 1 N H2SO4, and the absorbance values were measured at 450 nm using an ELISA reader. Anti-SARS-CoV-2 antibody (NIBSC code 20/130) was also included in the assay as a reference standard. The cut-off for the assays was set at twice of average optimum density (OD) value of negative control. The endpoint titre of a sample was defined as the reciprocal of the highest dilution that had reading above the cut-off value ^3, 24^.

### Neutralizing antibody evaluation

The plaque reduction neutralization assay (PRNT50) was performed against the B.1 (NIV2020-770, GISAID accession number: EPI_ISL_420545), Alpha [B.1.1.7, hCoV-19/India/20203522 SARS-CoV-2 (VOC) 20211012/01], Beta (B.1.351, NIV2021-893, GISAID accession number: EPI_ISL_2036294) and Delta (B.1.617.2, NIV2021-1916, GISAID accession number: EPI_ISL_2400521) strains which were isolated from clinical samples of COVID-19 positive samples at Maximum Containment Laboratory of ICMR-NIV, Pune.

The sera were serially diluted four-fold and mixed with an equal amount of B.1, Alpha, Beta and Delta virus suspension (50-60 Plaque forming units in 0.1 ml) separately. Post incubation at 37°C for 1 hr, each virus-diluted serum sample (0.1 ml) was inoculated onto duplicate wells of a 24-well tissue culture plate of Vero CCL-81 cells. After incubating the plate at 37°C for 1 hr, an overlay medium consisting of 2% Carboxymethyl cellulose (CMC) with 2% fetal calf serum (FCS) in 2× MEM was added to the cell monolayer. The plate was further incubated at 37°C with 5% CO2 for 5 days. At assay termination, plates were stained with 1% amido black for an hour. NAb titers were defined as the highest serum dilution that resulted in >50 (PRNT50) reduction in the number of plaques^25^.

### Phenotyping of lymphoid cells of lysed whole blood

To characterize generalized CMI, we analysed the phenotype and proportion of different lymphoid subsets (T helper, T cytotoxic, NK and B cells) in peripheral blood of each of the three study groups. 0.1 ml of well-mixed anti-coagulated whole blood was surface-stained with appropriate fluorochrome-conjugated antibodies (BD Biosciences). Two sets of sample tubes were prepared, one for T cells (CD3-PerCP, CD8-PE, CD4-FITC) and the other for B and NK cells (CD45-PerCP, CD3-FITC, CD16+56-PE, CD19-APC). After incubation for 30 min at 40C in dark, 2ml of lysis buffer was added to each tube, vortexed and incubated at room temperature for 12 minutes. 2 ml of washing solution was added to each tube. The sample tubes were centrifuged at 200x for 5 minutes and supernatant was aspirated carefully. The cell pellets were suspended in 500μl wash buffer and vortexed. Samples were acquired and analysed on Attune NxT flow cytometer using Attune NxT software (Thermo Fisher Scientific). (Figure-6)

#### Statistical analysis

The normality of the data was compared using the Shapiro-Wilk test for normality using two-tailed test in GraphPad Prism v 9.2.0. The ELISA titres against S1-RBD, N protein and inactivated SARS-CoV-2 (whole virus) was compared in sera of the three groups and analysed statistically using the Kruskal-Wallis test along with Dunns’ multiple comparisons in GraphPad Prism. Wilcoxon signed-ranks test was performed to analyse the statistical significance of the neutralization titre of the each individual sera against the different strains. Kruskal-Wallis test along with Dunns’ multiple comparisons was performed to assess the significance in NAb titres of the sera against each strain.

## Data Availability

All the data pertaining to this study is available with the Corresponding Author.

## Acknowledgement

We sincerely acknowledge the excellent support of Ms. Aasha Salunkhe, Mr. Chetan Patil, Dr. Rajlaxmi Jain, Mr. Prasad Sarkale, Mr. Shreekant Baradkar, Mr. Yash Joshi, during the study. We are thankful to Shri Amit Mohan Prasad, Additional Chief Secretary, Medical, Health and Family Welfare Department, Government of Uttar Pradesh for allowing us to carry out this study and extending all the required support.

## Author contributions

PDY, RKS and BB conceived the idea. PDY, RK and KZ designed the study. GD, KZ, RRS, RS and SC contributed patient recruitment and data acquisition. GS, HK, ASA, DAN, GRD carried out the experiment. PDY, RKS supervised all parts of the study, PDY, SK, RRS, HK performed analysis and PDY, NG, SK, PA, SP wrote the manuscript. All the authors have approved the manuscript.

